# Leveraging neural drive to assess hand motor function in individuals with chronic stroke

**DOI:** 10.1101/2025.05.30.25328498

**Authors:** Nicholas Tacca, Jackson T. Levine, Mary K. Heimann, Bryan R Schlink, Samuel C Colachis, Austin Bollinger, Collin Dunlap, Philip Putnam, Michael J Darrow, Lauren Wengerd, Josè L Pons, David A Friedenberg, Eric C Meyers

## Abstract

**Background:** Stroke is a leading cause of disability, with up to 80% of survivors experiencing motor impairments. These impairments are attributed to various factors, including reduced neural drive and altered motor unit firing patterns. Rehabilitation aims to restore motor function by enhancing motor unit recruitment and synchronization. High-density electromyography (HD-EMG) is a valuable tool for evaluating these changes in motor unit activity.

**Methods:** We tested a HD-EMG wearable forearm sleeve to investigate the relationship between motor function and motor unit properties including firing rate, motor unit module activation, and coherence. Seven individuals with chronic stroke and seven able-bodied individuals attempted 12 controlled hand and wrist movements while EMG was recorded. Motor units were decomposed across all movements using convolutive blind source separation.

**Results:** Fewer motor units were detectable in individuals with stroke compared to able-bodied participants. There was a significant reduction in motor unit firing rate during specific movements such as wrist flexion and hand open. Motor unit coupling and activation were altered following stroke, with reduced module activation in 8 of the 12 movements attempted. Furthermore, a reduction in coherence for gross movements and an increase in coherence for more dexterous thumb movements suggest altered neural drive to motor units after stroke that is differentially tuned to the complexity of movement. A combined neural control signature, consisting of multiple motor unit features, demonstrated strong correlation (***R*^2^ = 0.81**) with clinical motor function scores.

**Conclusions:** This study demonstrates that HD-EMG can capture detailed motor unit activity and neural control characteristics across multiple forearm muscles in individuals with chronic stroke. By integrating multiple HD-EMG features, this approach provides new insights into neuromuscular alterations linked to hand motor function after stroke. These findings support the use of HD-EMG for monitoring recovery, predicting outcomes, and guiding more targeted rehabilitation, thus advancing both stroke research and patient care.

## 1 Background

Stroke is a leading cause of disability worldwide, with more than 12 million new cases reported globally and over 795,000 cases annually in the United States alone [1, 2]. Approximately 80% of stroke survivors experience motor function impairment, which significantly impacts their quality of life [3]. This impairment is believed to be attributed to a complex interplay of factors, including reduced neural drive, muscle atrophy, disorganized motor unit firing patterns, and changes in coordinated motor control [4–6]. These alterations are particularly pronounced in the upper extremity, resulting in diminished force generation, reduced speed, loss of independent joint control, and diminished dexterity during voluntary movements [5]. Impairments to hand motor control can be significantly disabling because they affect an individual’s ability to perform essential daily tasks that require dexterity and fine motor skills [3]. Understanding these changes is crucial for monitoring rehabilitation progress and developing effective therapeutic strategies aimed at restoring motor function.

Motor units are the fundamental elements of muscle contraction, responsible for translating neural signals into mechanical output [7]. They are essential for the precise control of muscle force and coordination. Advances in high-density surface electromyography (HD-EMG) technology and algorithms have made it feasible to non-invasively decompose motor units during isometric contractions [8–12]. Recently, several studies have demonstrated the ability to decompose motor units during controlled dynamic movements [13–20]. This allows for the assessment of changes in motor unit activity post-stroke [5, 6, 21] during more realistic and functional tasks, facilitating its use in both clinical settings and home-based rehabilitation. Monitoring changes in motor unit activity in the hand offers a promising way to detect subtle neural recovery or maladaptive compensation during upper extremity rehabilitation. This information could help occupational therapists and neurorehabilitation clinicians adjust therapy plans in real-time to maximize function. In particular, motor units involved in dexterous movement of the hand exhibit a high degree of flexibility and may be used as highly sensitive biomarkers [22]. These insights may help tailor treatment plans to each person’s specific needs, leading to more personalized and effective rehabilitation strategies [23, 24]. A user-friendly HD-EMG sleeve could provide therapists with realtime, objective insight into motor control changes, guiding intervention decisions such as task selection, intensity progression, or device use [25].

By examining specific characteristics of motor unit firing, we can gain deeper insights into motor control mechanisms. Previous studies have identified key properties of altered motor unit activity post-stroke, which include firing rate[26], recruitment [6, 21], coupled activity, and discharge coherence [4], each providing unique insights into specific aspects of motor control. The rate at which motor neurons discharge action potentials is indicative of the neural drive to muscles [7, 27, 28]. Reduced motor unit firing rates have been observed in individuals post-stroke, which correlate with impaired motor function [5, 26, 29] and may contribute to muscle weakness characteristically experienced after stroke.

In addition to assessing motor unit firing rate, it is also useful to examine the coupling of motor units and the coordinated activation of groups of motor units [28, 30–34]. Traditionally, the central nervous system (CNS) is believed to control movements by activating groups of muscles, or modules, allowing for reduction in the complexity of neural commands sent from the CNS [35–37]. In an effort to quantify the source of the loss of independent joint control post-stroke, muscle modules (or synergies) have been found to be impaired in their organization, with the altered organization relating to functional impairment [38, 39]. The recent extension of this analysis technique to the motor unit level [40] provides opportunity to investigate if the coordinated activation of groups of motor units, motor unit module activation, is also altered post-stroke. We hypothesize that the abnormal coordination of muscles, which results in the loss of independent joint control, will also be apparent at the motor unit level and contribute to functional impairment observed after a stroke.

Additionally, motor unit coherence analysis extends the understanding of motor unit coupling by assessing the degree of common input that motor units receive from various sources [27, 28]. Motor units integrate information from these various inputs to facilitate the modulation of muscle force during contractions [27, 28]. Different frequency bands provide insight into specific sources of neural communication received by the pool of motor units: the delta/theta band (0-8 Hz) is related to neural drive and force development [27]; the alpha band (8-12 Hz) is associated with peripheral afferent feedback and pathological tremor [41]; and the beta (12-30 Hz) and gamma (30-120 Hz) bands are linked to supraspinal centers [42–44]. By analyzing these coherence patterns, we can gain a deeper understanding of the neural strategies employed by the CNS to control movement, particularly in the context of stroke rehabilitation. Furthermore, there is evidence that the dominant hand, which tends to exhibit greater dexterity, has reduced common synaptic inputs, characterized by a reduction in motor unit coherence within and between muscles of the hand [45]. We hypothesize that following a stroke, individuals will exhibit increased motor unit coherence during dexterous movements compared to able-bodied individuals due to the damage to the corticospinal pathways primarily responsible for dexterous control and compensation of more grossly activated pathways (e.g., reticulospinal)[46], which may increase synchronization of motor units. While each feature provides valuable insights into different aspects of motor control, combining them into a comprehensive metric may offer a more encompassing and interpretable signature of motor function [47]. This integrated approach can better elucidate the neural strategies employed by the CNS to control movement, particularly in the context of stroke rehabilitation.

In this study, we employ a wearable HD-EMG sleeve that enables the simultaneous recording of motor activity across multiple forearm muscles during controlled dynamic movements. This technology offers ease of use with a streamlined setup, making it suitable for clinical environments and at-home use. Our focus on hand movements in individuals with stroke addresses relevant motor function for activities of daily living. By decomposing motor units and analyzing various firing characteristics during attempted movements, we aim to comprehensively characterize neural alterations post-stroke. Furthermore, by integrating four motor unit features, we developed a comprehensive neural control signature that correlates strongly with clinical motor function scores. This approach elucidates neuromuscular alterations and has the potential to enable more targeted rehabilitation interventions. By leveraging a user-friendly wearable system, this work demonstrates the potential for translating non-invasive wearable neurotechnology to a clinical setting to improve motor rehabilitation for individuals with stroke.

## 2 Methods

### 2.1 Study participants

The study was conducted at Battelle Memorial Institute in accordance with Battelle’s Institutional Review Board (IRB0779 and IRB0773). Informed consent was provided by the participants following the standards outlined in the Declaration of Helsinki. EMG was recorded from the affected arm of individuals with a history of stroke and hemiparesis (N=7, 3 female, 4 male; mean age 60±5 years) and able-bodied individuals (N=7; mean age 27±1 years). Full inclusion and exclusion criteria can be found in the Supplementary Information. Prior to EMG data collection, all individuals poststroke underwent standardized clinical assessments, including the voluntary movement components of the upper extremity section of the Fugl-Meyer (UEFM) [48], the Box and Blocks test, and the Modified Ashworth Scale (MAS) to evaluate gross arm and hand coordination, as well as finger and wrist spasticity. Full clinical assessment results are contained in Table 1.

**Table 1.**
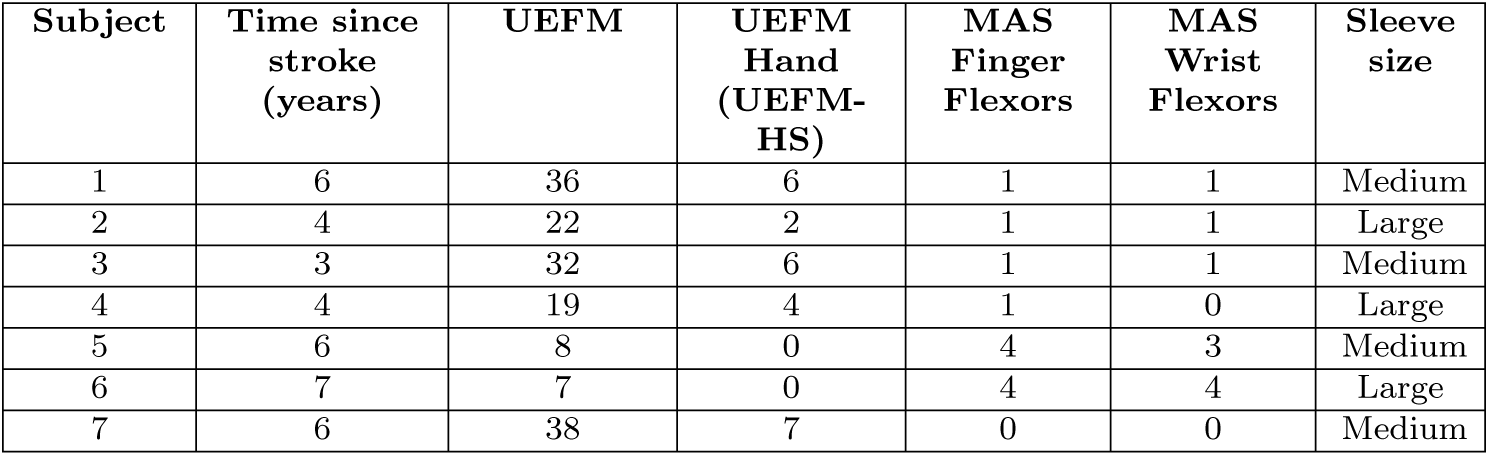
Clinical assessments and sleeve size for individuals with stroke. UEFM: upper-extremity Fugl-Meyer Score; total voluntary movement score ranges from 0-60 [48], with higher scores indicating less severe impairment. UEFM-HS is the hand subscore of the UEFM, ranging from 0-14. MAS: Modified Ashworth Scale; each joint is scored individually from 0-4, where higher scores indicate greater spasticity.

| Subject | Time since stroke (years) | UEFM | UEFM Hand (UEFM-HS) | MAS Finger Flexors | MAS Wrist Flexors | Sleeve size |
| --- | --- | --- | --- | --- | --- | --- |
| 1 | 6 | 36 | 6 | 1 | 1 | Medium |
| 2 | 4 | 22 | 2 | 1 | 1 | Large |
| 3 | 3 | 32 | 6 | 1 | 1 | Medium |
| 4 | 4 | 19 | 4 | 1 | 0 | Large |
| 5 | 6 | 8 | 0 | 4 | 3 | Medium |
| 6 | 7 | 7 | 0 | 4 | 4 | Large |
| 7 | 6 | 38 | 7 | 0 | 0 | Medium |

### 2.2 Experimental setup

This study used the same dataset as our previous studies [47, 49]. A depiction of the experiment overview is shown in Figure 1. Participants were seated comfortably at a table with a computer monitor in front of them. The NeuroLife^®^ EMG sleeve array was used to collect EMG data from the forearm as participants attempted various cued hand and wrist movements (Figure 1A). The EMG sleeve consists of a highdensity array of electrodes embedded in a stretchable fabric with up to 75 EMG channel pairs. Prior to donning the sleeve, participants’ forearms were sprayed with an electrode solution spray (Signaspray, Parker Laboratories). Individuals post-stroke wore the sleeve on the affected arm while able-bodied participants wore the sleeve on their dominant arm. Surface EMG was recorded using a bipolar configuration. Sleeve size for each participant was determined based on comfort and fit (Table 1).

**Fig. 1.**
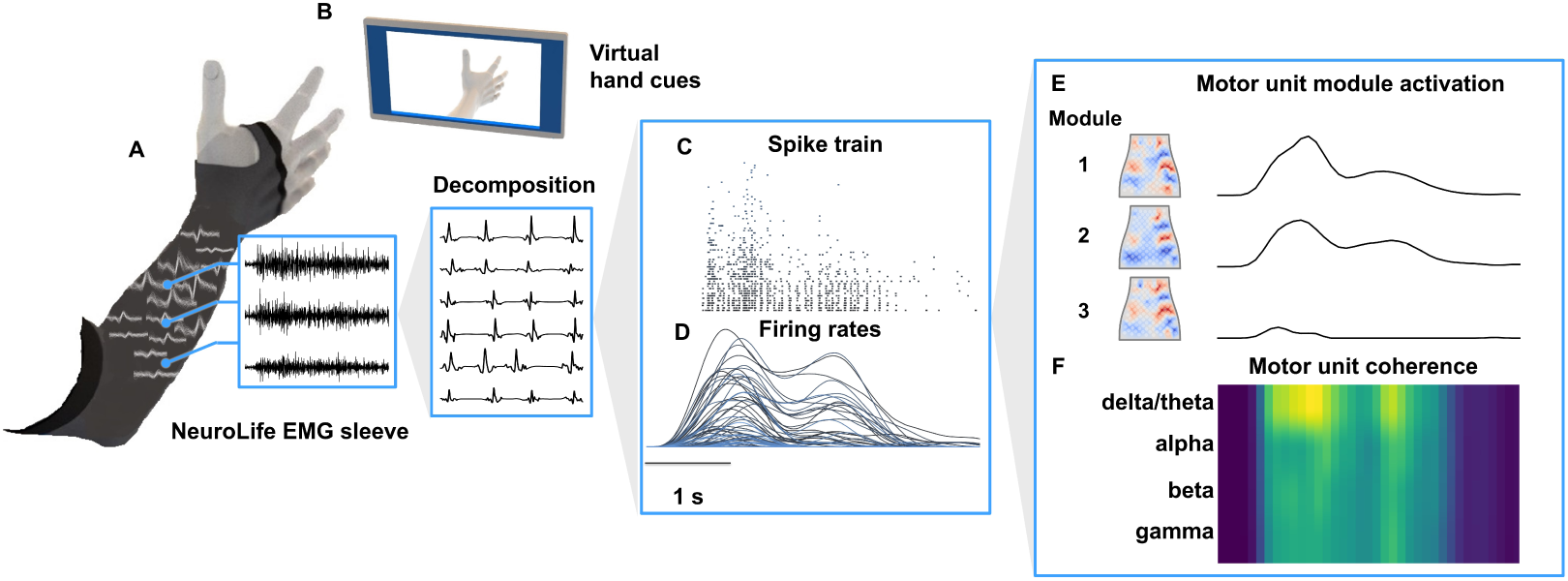
Experiment overview. **A.** The NeuroLife EMG sleeve array records high-density EMG from the forearm with up to 75 bipolar EMG channels. Representative motor unit action potential waveforms extracted from the spike discharge timings are depicted for one motor unit across channels on the sleeve. The EMG signal consists of the summation of individual motor unit action potentials and noise, which can be decomposed using convolutive blind source separation techniques. Different motor unit features (**D-F**) were extracted from firing patterns and correlated with clinical scores to show the feasibility of predicting motor function. **B.** Computer monitor that displays hand movement cues via a virtual hand for participants to attempt. **C.** Exemplary motor unit spike train decomposed from a hand open attempt by a subject in the stroke group. **D.** Subsequent motor unit firing rates smoothed with a 300 *ms* hamming window. **E.** Motor unit modules decomposed from motor unit firing rates using non-negative matrix factorization mapped to the flattened sleeve array (left) and module activation (right) during the hand open attempt. **F.** Global motor unit firing coherence between random cumulative spike trains made up of two motor units computed across the delta/theta (0-12*Hz*), alpha (8-12*Hz*), beta (12-30*Hz*), and gamma (30-120*Hz*) bands during the hand open attempt.

A virtual hand on the monitor displayed various hand, wrist, and forearm movements for participants to attempt (Figure 1B). A licensed occupational therapist scored each movement using the Action Research Arm Test (ARAT) [50] scoring scheme (0 = no movement; 1 = incomplete range of motion; 2 = complete range of motion but impaired; 3 = normal). This ensured scoring consistency while aligning with how therapists assess movement quality in practice. Supplementary Figure 1 shows the observed movement scores across all attempted movements. Both stroke and able-bodied participants attempted the following twelve movements during a single session: Hand Close, Hand Open, Pointing Index, Thumb Flexion, Thumb Extension, Thumb Abduction, Wrist Supination, Wrist Pronation, Wrist Flexion, Wrist Extension, Thumb Two Point Pinch, and Key Pinch. EMG signals were recorded in structured blocks, each block including five repetitions of single movements. Blocks consisted of alternating phases of rest and the cued movement. Each cue was prompted for 4-6 seconds for individuals with stroke and 2-3 seconds for able-bodied participants. The first five movement attempts from each participant were used to have an equal number of movement attempts across participants and subject groups.

### 2.3 Neural drive feature extraction

#### 2.3.1 Preprocessing and HD-EMG decomposition

An Intan Recording Controller was used to sample EMG data at a rate of 3,000 *Hz*. The recorded EMG data was subsequently filtered using a 10*^th^* order Butterworth filter between 20 and 400 *Hz* and a notch filter at 60 *Hz*. The open-source motor unit decomposition package [51] based on work by Negro et al. [8] was used to extract motor unit firings from the filtered EMG signal offline across all movements. Default parameters were used except where noted below to optimize decomposition for the sleeve array, particularly due to the larger interelectrode distance (18.5-25 *mm*) and size of the electrodes (12 *mm*). An extension factor of 14 was used for all subjects regardless of sleeve size. This was chosen based on the recommendation from Negro et al. [8] (1000*/n_channels_*) for the medium size sleeve configuration (70 channels). We fixed the extension factor across sleeve sizes for simplicity, since results from motor unit decomposition are not substantially influenced by the extension factor within the range of 8-31 for surface EMG [8]. A silhouette threshold of 0.90 was used based on [8] to only keep reliable sources. To ensure motor units were not redundant, two motor unit spike trains whose firings coincided for at least 20% of the spike times were considered duplicates. If a pair of motor units exceeded this threshold, then one of these motor units was removed. The minimum interspike interval was set to 3 *ms* to include spikes from short burst motor neurons in addition to motor neurons involved in sustained muscle contractions in the range of 10 - 40 *Hz* [52–54]. No maximum interspike interval was set.

Since motor units were extracted from a concatenated dataset of movements, decomposition was performed in batches using the built-in EMG decomposition package GPU capabilities. The batch size consisted of 36,000 samples (12*s*) to include at least one full movement attempt and rest period per batch. In order to not preferentially find motor units for movements attempted at the beginning of the dataset, batches were randomly shuffled from the full concatenated dataset. Since the decomposition algorithm depends on a random peak initialization, the full dataset of batches was sampled twice in random order to ensure motor units were extracted across all movements sufficiently. As the separation source filters (*B* matrix from [8]) were found batch by batch, the previous sources were saved, and new sources were decomposed on subsequent batches. To ensure the appropriate number of sources were extracted and to increase decomposition efficiency, the maximum number of sources parameter that set how many sources the algorithm was searching for was iteratively updated in each batch as new sources were found. The initial value for the maximum number of sources was set to half the number of EMG channels.

#### 2.3.2 Motor unit post-processing

Following decomposition, motor units and their corresponding spike trains (Figure 1C) were processed further to increase reliability and repeatability based on the following work [55]. First, sources that were likely noise were removed based on a hyperspiking criteria. If a motor unit source fired on average more often during the rest periods than movement-selective periods, the source was considered to be hyperspiking, and most likely responded to artifact rather than muscle contraction. Next, errant spikes were removed corresponding to artifacts in the data arising from changes in electrode contact and physical interaction with the electrodes. Due to the introduction of signal discontinuities from concatenation of movement blocks, if more than 80% of motor units fired at any given time, these spikes were removed.

While the HD-EMG sleeve provides high-resolution information across and within muscles, it is important to identify the muscles that are innervated by the decomposed motor units to provide functional relevance. Understanding which muscle group a motor unit corresponds to can help evaluate the common synaptic input through both local and global firing characteristics as well as identify potential focal weaknesses or sites of impairment. Therefore, a mapping procedure was used to assign motor units to flexor or extensor muscle groups. First, motor units were assigned to movement classes based on how often they fired during a cue. Hand Open and Wrist Extension were used to determine the extensor movements, while Hand Close and Wrist Flexion were used to determine the flexor movements. To be assigned to a class, a motor unit needed to fire with an average firing rate of at least 2 *Hz* during the cues [56].

Due to joint stabilization through co-contraction, motor units could be assigned to both flexors and extensors based on movement attempt alone. Therefore, an additional spatial mapping procedure was conducted to determine the corresponding muscle groups for motor units. Motor unit action potential amplitude maps were created to assess motor unit amplitude localization on the sleeve. Waveforms were extracted for each motor unit via a spike triggered average in which 30 *ms* windows of EMG data centered about discharge impulses were summed. The waveforms were further smoothed by taking the average with the surrounding neighboring channels (Figure 2A). Amplitudes at different time points throughout the waveform were extracted to create amplitude maps (Figure 2B).

**Fig. 2.**
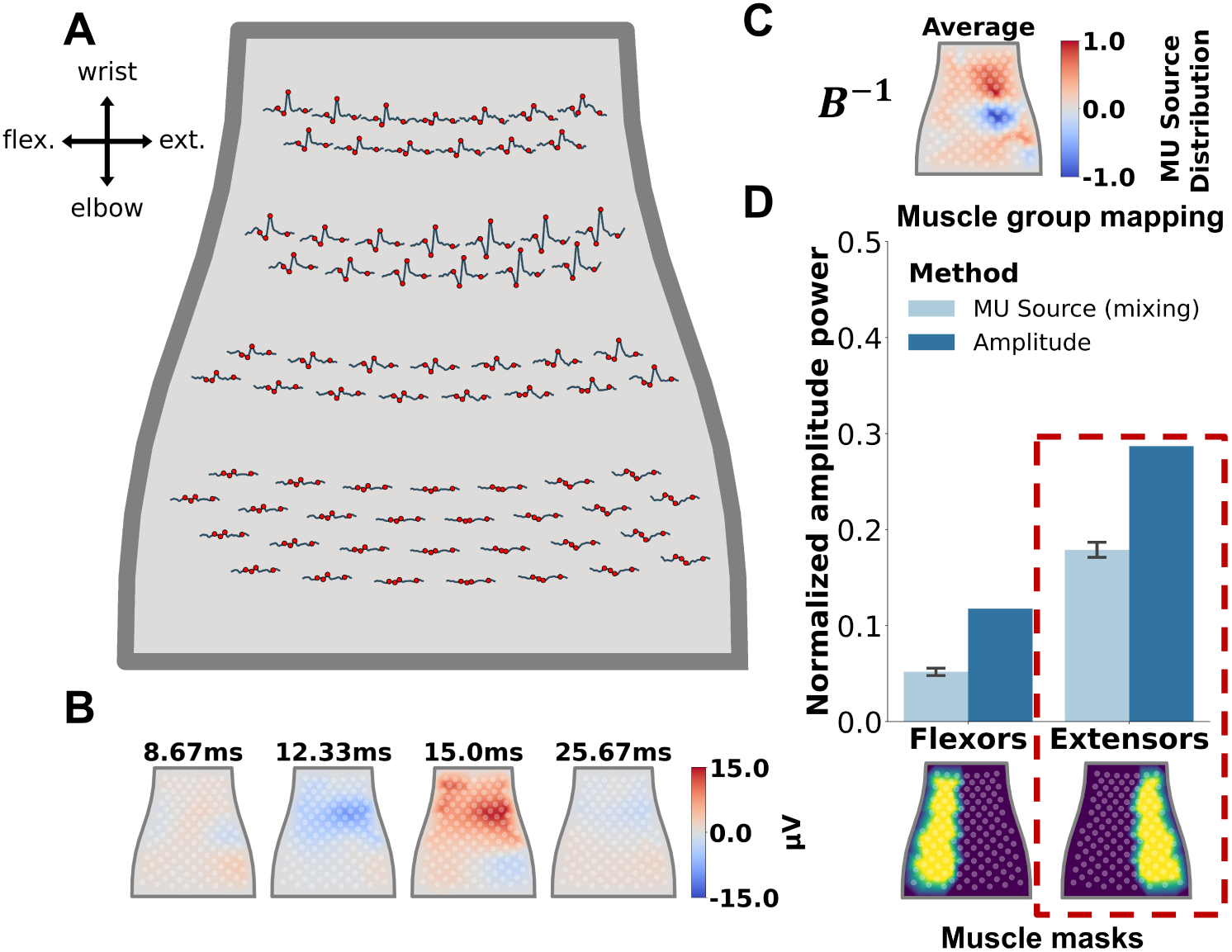
Mapping motor units to muscle groups. **A.** Motor unit action potential waveforms for an extensor motor unit plotted on the sleeve array across the respective channels. Waveforms were generated by taking a spike triggered average of the filtered EMG signal at the discharge sample. Red markers represent time points of interest along the waveform. **B.** Amplitude maps of the spike triggered average waveforms at the time points shown in **A**. **C.** Inverse blind source separation (mixing) filter averaged over extensions representing MU source distribution. **D.** Normalized peak amplitude (t=15.0 *ms*) and MU power source distribution (averaged over extensions *±* SEM) mapped to flexors and extensors. Below are flexor and extensor masks used for mapping motor units to muscle groups.

Furthermore, to understand the physiological weighting of motor unit source activity, the mixing matrix (*B^−^*^1^) from blind source separation was used to visualize motor unit source power on the EMG sleeve (Figure 2C) based on Equation 1:

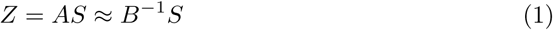

Here, *Z* refers to the extended and whitened EMG data with shape *n_ext channels_ × n_samples_*. *A* is the mixing matrix (*n_ext channels_× n_sources_*) that reconstructs the extended and whitened data from the separated sources *S* (*n_sources_ × n_samples_*). When the sources are squared, the resulting peaks represent motor unit impulses. *B* denotes the unmixing filter (*n_sources_ × n_ext channels_*) computed using a fixed point algorithm [8] that, when multiplied with the extended and whitened data, extracts independent motor unit sources. We assume then that *B^−^*^1^ is a good approximation for the mixing matrix *A*. Thus, we can visualize individual extensions and the average over the extensions from *B^−^*^1^ mapped onto the sleeve array to understand how the motor units are weighted to individual channels (Figure 2C). Similarly, we can use methods from our previous work [47] to map EMG channels to muscle groups for further physiological interpretation of motor unit weighting. In this work, we also use a simplified mapping to flexors and extensors to understand the weighting of motor units to muscle groups. When the sleeve is donned on the right arm, flexor channels are on the left half of the sleeve, whereas extensor channels are on the right half of the sleeve (Figure 2D).

These masks were used to map both the amplitude maps and average source weighting to the corresponding muscle groups. Taking into consideration the amplitude power at t = 15 *ms* and mixing matrix mapping (Figure 2D), a final muscle group was determined by taking the average ratio between muscle groups that yielded greater than 50% selectivity. If we were unable to determine whether a motor unit corresponded to flexors or extensors based on firing and spatial mapping ratio, this motor unit was removed from all analyses. We recognize that this classification potentially excludes motor units from muscles that perform other functions at the wrist (e.g., pronation, supination), but we selected this approach for simplicity of the analysis and availability of sufficient number of motor units.

Lastly, all motor units and spike trains were visually assessed to verify the remaining motor units were valid [55]. Motor unit action potential waveforms were visualized by taking the average over the maximum source power from the mixing matrix over the top 10 channels with maximum response. Waveforms were visually inspected to ensure the motor unit represented a high signal-to-noise source. Any waveforms with distorted action potential shapes, multiple noisy traces, or minimal consistency between waveform traces were removed. This provided confidence that all sources for subsequent analyses were likely motor unit sources.

#### 2.3.3 Motor unit firing rate and recruitment duration

Once motor unit post-processing was finalized, instantaneous firing rates for each motor unit were determined by taking the inverse of the interspike-interval. The instantaneous firing rates were averaged over 100*ms* bins to both smooth and align firing rates with the presented cues (Figure 1D). To assess firing rate by movement, each movement repetition was epoched at the start of the shifted cue to align with physical movement attempt. For individuals with stroke, cues were shifted by 600 *ms*, whereas for able-bodied subjects, cues were only shifted for 300 *ms* to account for quicker reaction times. The first two seconds of each shifted cue were used for comparisons between groups. Average motor unit firing rates were compared between groups by movement. To assess the changes in the motor activation and sustained muscle contractions post-stroke, motor unit recruitment and de-recruitment durations were calculated for both able-bodied and stroke populations. Motor unit recruitment period was determined by taking the time to reach maximum firing rate from the initial onset of motor unit activity during each movement attempt. The de-recruitment period consisted of assessing the ability to sustain contractions by taking the time from the maximum firing rate to minimum firing rate during a movement attempt. A conservative minimum firing rate threshold at recruitment of 2 *Hz* was used to only select active motor units during movement attempts [56].

#### 2.3.4 Motor unit modules

To extract pools of highly coupled motor units, non-negative matrix factorization (NMF) from the Scikit learn toolbox [57] was used to reduce the dimensionality of the smoothed motor unit firing rates (*X*: *n_sources_ × n_samples_*) across all movements into time-invariant motor unit modules (*W* : *n_sources_ × n_modules_*) and time-varying motor unit module activations (*H*: *n_modules_ × n_samples_*) (Figure 1E) based on Equation 2.

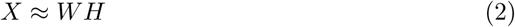

NMF was chosen based on previous studies assessing motor unit modules [32, 58] and has been shown to be robust across datasets when extracting muscle synergies [59]. The number of modules required to fully explain the common synaptic input to motor units for an individual was determined based on an 85% variance accounted for (VAF) threshold between the original firing rates and reconstructed firing rates [38, 47, 59]. When comparing module activation between subjects and subject groups, the peak module activation was taken during the attempted movement across the most active module. Motor units were considered to be in a module if the motor unit weighting was greater than zero. Additionally, motor units were not constrained to be in a single module. The ratio between number of motor units in a module over total number of motor units decomposed for each subject was used to understand the relative contribution of motor units to individual modules. To visualize the weighting of modules mapped to EMG channels, the modules were multiplied with the mixing matrix calculated from motor unit decomposition (Equation 3).

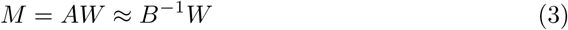

*M* is the motor unit module matrix, which maps the whitened and extended data to the motor unit modules (*n_ext channels_ × n_modules_*). Therefore, each extension or the average over all extensions can be visualized on the sleeve array to understand channel weighting of the module. As in the case with visualizing amplitude maps and motor unit source power, the module source power was mapped to muscle groups for further physiological interpretation.

#### 2.3.5 Motor unit coherence

While motor unit modules extract highly coupled motor units, it may be beneficial to assess both a local and global firing coherence between motor units within and across muscle groups, respectively (Figure 1F). Therefore, we conducted four different motor unit coherence analyses, namely intra-flexor, intra-extensor, inter-flexor-extensor, and global motor unit coherence. Methods were consistent between analyses except for which motor units were selected per group. First, a pair of cumulative spike trains (CSTs) of two motor units were computed per subject by randomly selecting unique motor units and summing their binary discharges for a total of 100 random permutation comparisons, similar to previously described methods [60]. Subjects 4 and 6 were excluded from the intra-flexor and intra-extensor analysis, respectively, as well as the inter-flexor-extensor analysis, due to having less than four motor units per muscle group. Squared coherence between CSTs was computed over time from 200 *ms* bins (600 samples) with 50% overlap (coherence sample every 100 *ms*). Within each bin, windows of 512 samples (171 *ms*) with 95% overlap between 0 and 120 *Hz* were used to calculate coherence (*n_bins_ × n_CST s_ × n_CST s_× n_frequencies_*) where *n_CST s_* = 2 for a comparison [61]. In total, coherence was computed over 20 equally spaced frequencies within the specified range. Coherence matrices were then Z-transformed and reduced to four frequency bands by taking the average over frequencies within each band (delta/theta: 0-8 *Hz*, alpha: 8-12 *Hz*, beta: 12-30 *Hz*, and gamma: 30-120 *Hz*) [62]. Following this, the unique comparison (upper right value) of the coherence matrices was returned for each frequency band (*n_bins_ ×n_bands_*). For the inter-flexor-extensor analysis, only CST pairs between muscle groups were assessed. After 100 random pairwise permutations, coherence was averaged over comparisons. Finally, the resulting coherence was smoothed over time using a 500 *ms* hamming window. Average peak Z-coherence during the cue was compared between groups for individual movements.

### 2.4 Clinical correlations

To assess whether the common drive to motor units could be used to predict motor function, features presented in this work, namely motor unit firing rate, motor unit module activation, motor unit module ratio, and global motor unit coherence in the delta/theta band, were correlated with UEFM-HS and MAS. Features were correlated with clinical scores by movement as applicable. Within and across muscle specific motor unit coherence were not considered since not enough motor units were decomposed per muscle group for two participants with stroke (subject 4: flexors, subject 6: extensors). The delta/theta band was selected over the other bands as values significantly differed between able-bodied individuals and individuals post-stroke. Select movements with high correlation were subsequently used to create a combined neural control signature. A concatenated feature set consisting of all the aforementioned features for Wrist Supination, Wrist Pronation, and Wrist Flexion were selected and reduced using principal component analysis. The first principal component was then correlated with UEFM-HS to demonstrate the proposed efficacy of the neural control signature (Figure 1G). Loadings were visualized to assess the contribution of each feature to the first principal component.

### 2.5 Statistical analysis

We used Lilliefors tests to test normality of distributions. For normal distributions, unpaired t-tests were used to compare independent groups (Figures 3E, 5G, 6C). The Kruskal-Wallis H-test was used for non-normal distributions (Figures 4B-C, 5E). A p-value of 0.05 was used for single comparisons. For multiple comparisons (Figures 4B-C, 5E, and 6C), p-values were adjusted using a one-step bonferroni correction. All statistical analyses were performed using Python 3.8 using SciPy [63] and statsmodels [64]. For all figures, * indicates *p <* 0.05, ** indicates *p <* 0.01, and *** indicates *p <* 0.001. Error bars in figures indicate mean *±* standard error of the mean (SEM).

**Fig. 3.**
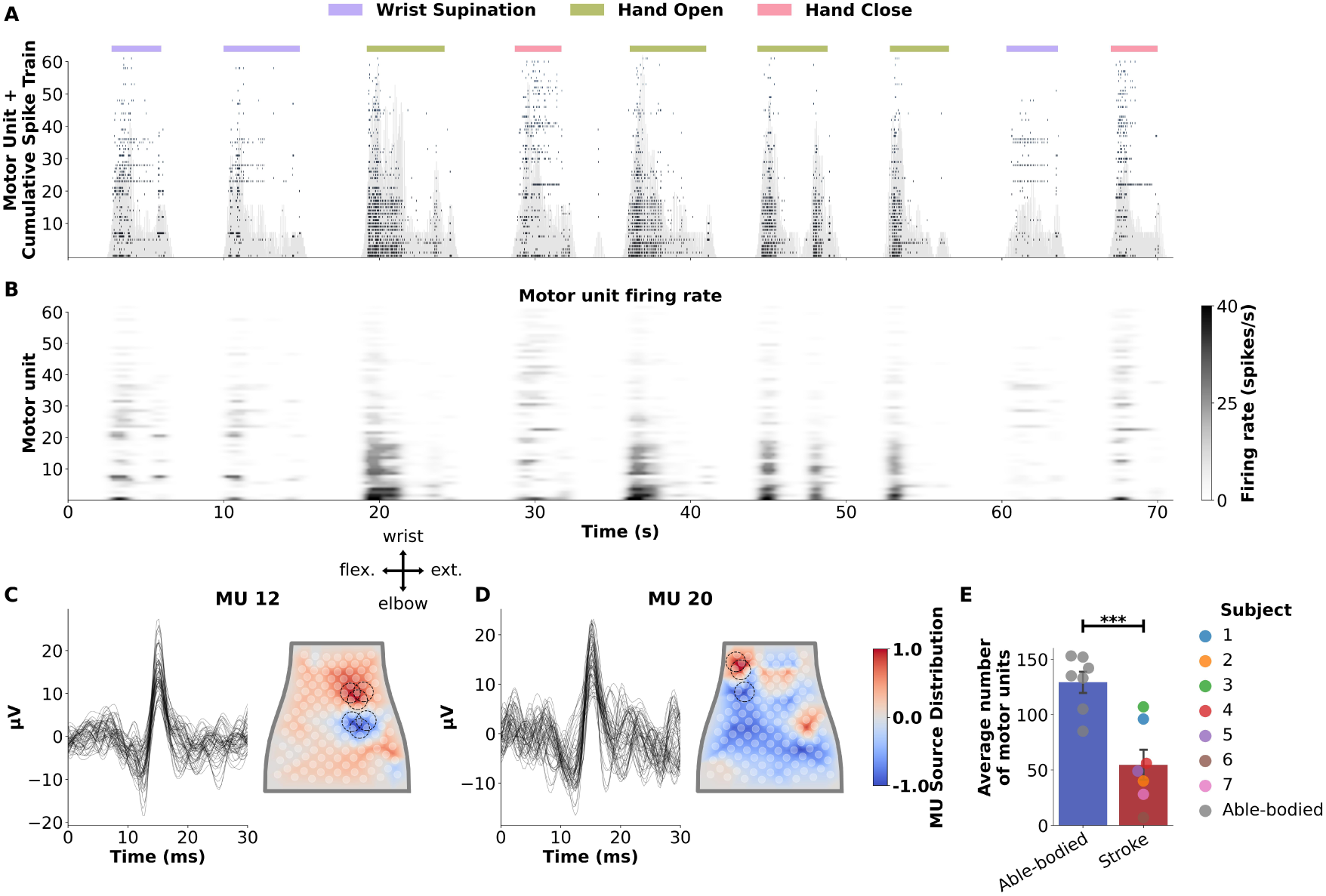
Motor unit decomposition using a wearable sleeve. **A.** MU spike train decomposed from a stroke participant attempting three movements. MUs are sorted based on the number of firing impulses. The cumulative spike train (CST) smoothed with a one second hamming window is shaded in light gray behind the spike train. **B.** Smoothed motor unit firing rate is shown below for the respective motor units. Firing rates were smoothed with a 300 *ms* hamming window. **C-D.** MU action potential waveform (left) and source distribution (right) for two representative MUs. Waveforms were extracted by taking a spike triggered average from the filtered EMG signal centered about each discharge impulse. Waveforms were subsequently averaged across the channels circled on the heatmap for a single action potential visualization. The MU spatial source distribution is represented by the inverse blind source separation filter averaged over the 14 extensions. Heatmaps were normalized between -1 and 1 and clipped with a 99% threshold to spatially localize MU source power on the sleeve array. **E.** Average number of motor units extracted across all 12 movements for able-bodied and stroke participants.

**Fig. 4.**
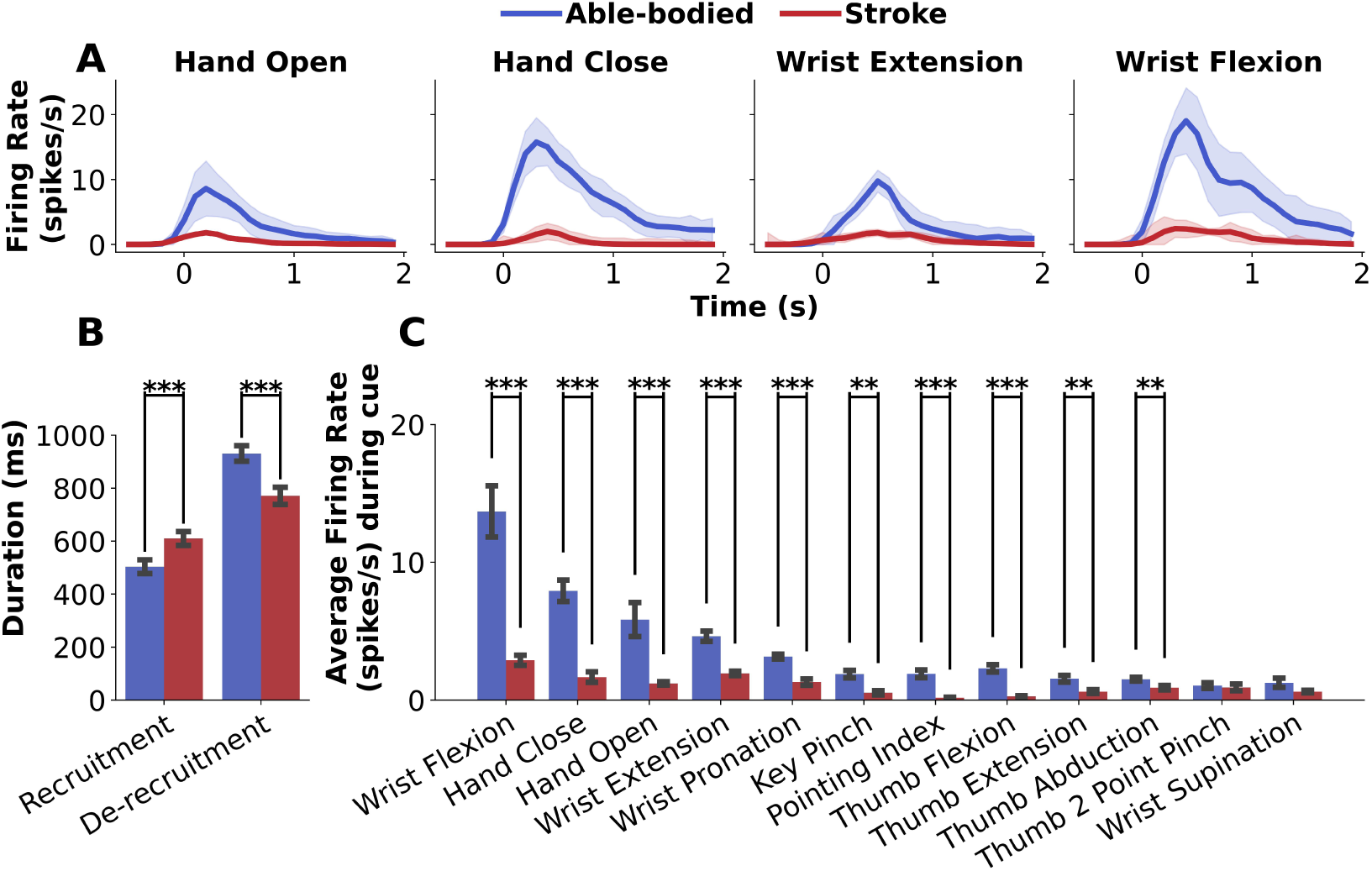
Motor unit firing rates are affected after stroke during find hand movements. **A.** Average firing rate *±* standard error of the mean across all subjects over Hand Open, Hand Close, Wrist Extension, and Wrist Flexion movement attempts. **B.** Average motor unit recruitment and de-recruitment duration across all 12 movements. Individuals with stroke had a slower recruitment period and were not able to sustain muscle contractions for as long as able-bodied subjects. **C.** Average firing rate during cue. Stroke subjects had significantly reduced firing rate throughout 10 of the 12 movements.

## 3 Results

### 3.1 Detection of motor units in individuals with stroke using a wearable sleeve

Motor unit spike trains were decomposed for all subjects across all 12 movements attempted. Figure 3A shows an example spike train from a participant with stroke (subject 1) over multiple movement attempts. Noticeable patterns of firing are seen for the different movements. The CST across all motor units is shown in light gray beneath the spike train to provide a relative neural drive magnitude of global muscle innervation. Figure 3B shows the corresponding time-series motor unit firing rate heatmap. Motor units fired at a maximum rate of 40 *Hz* during hand open attempts, whereas firing rate was minimal during wrist supination attempts.

To determine whether decomposed motor units were valid, motor unit action potential waveforms were plotted by taking the spike-triggered average of the filtered EMG signal at impulse discharge times (Figure 3C-D). Sources with distorted waveforms were removed from the analysis, resulting in inclusion of only motor units with clean waveforms resembling an action potential shape. To localize waveforms on the sleeve, the mixing filters were used to select channels with highest source weighting. The average source weighting over extensions is shown mapped to the flattened sleeve on the right of the waveforms in Figure 3C-D. The motor unit depicted in **C.** (MU 12) corresponds to the extensor muscle group and is active during the hand open attempts. The motor unit depicted in **D.** (MU 20) is weighted to the flexor muscle group and is subsequently more active during hand close attempts.

An average of 55 *±* 14 motor units were detectable in the stroke group, with 129 *±* 10 detected in the able-bodied group (Figure 3E). Despite attempting the same movements, significantly more motor units were decomposed in the able-bodied group (*p* = 7.2 *×* 10*^−^*^4^).

### 3.2 Reduced motor unit firing rates during fine hand movements post-stroke

To assess whether the wearable sleeve could detect any changes in motor unit firing rate in the forearm while participants attempted dexterous hand movements, average motor unit firing rate by movement was compared between able-bodied and stroke groups (Figure 4). Average motor unit activation profiles varied between groups with the able-bodied group tending to have sharper increase in recruitment to a higher maximum firing rate and long de-recruitment periods (Figure 4A). On the other hand, the stroke group tended to have a more gradual motor unit recruitment period to a lower maximum firing rate, with the inability to sustain contraction for as long as able-bodied subjects.

To assess whether there were significant differences in initial motor activation and sustained muscle contraction, average motor unit recruitment and de-recruitment periods were compared between populations (Figure 4B). There was an increase in time to maximum firing rate in individuals with stroke (610 *±* 26 *ms*) compared to able-bodied individuals (504 *±* 26 *ms*) (*p* = 5.78*×*10*^−^*^7^). Additionally, the ability to sustain muscle contractions after reaching peak firing rate was affected post-stroke with stroke individuals having a shorter de-recruitment period from maximum to minimum firing rate (771 *±* 32 *ms*) compared to able-bodied individuals (931 *±* 29 *ms*) (*p* = 9.43 *×* 10*^−^*^4^).

The average firing rate during a cue was compared between groups to assess changes in muscle innervation following a stroke (Figure 4C). Motor unit firing rates were significantly reduced after stroke for 10 of the 12 movements attempted (Wrist Flexion: *p* = 6.06 *×* 10*^−^*^5^, Hand Close: *p* = 2.96 *×* 10*^−^*^9^, Hand Open: *p* = 1.43 *×* 10*^−^*^5^, Wrist Extension: *p* = 3.10 *×* 10*^−^*^6^, Wrist Pronation: *p* = 8.32 *×* 10*^−^*^4^, Key Pinch: *p* = 2.01 *×* 10*^−^*^3^, Pointing Index: *p* = 1.08 *×* 10*^−^*^5^, Thumb Flexion: *p* = 1.62 *×* 10*^−^*^8^, Thumb Extension: *p* = 4.25 *×* 10*^−^*^3^, and Thumb Abduction: *p* = 8.53 *×* 10*^−^*^3^).

### 3.3 Coupled motor unit grouping and activation is altered following stroke

Motor unit modules were extracted from motor unit firing rates using NMF to assess changes in population level motor unit activation by the CNS. Figure 5A-D shows results from a single motor unit module decomposed from a stroke subject’s (subject 4) motor unit firing activity. Time-invariant weightings to individual motor units in the module were extracted from the ***W*** matrix from NMF (Figure 5A). To visualize the module weighted to channels on the sleeve array and muscle groups (Figure 2C), the mixing matrix ***B^−^*^1^** was multiplied with the weighting matrix for each motor unit (Figure 5B) determined from the convolutive blind source separation motor unit decomposition algorithm. In this example, the highest weighted motor units are shown to visualize the dominant motor unit channel weighting of the module. However, all motor units with weighting greater than zero were considered to be included in the module to account for subtle firing characteristics. This particular module has strong dipoles in the lower left region of the sleeve predominantly corresponding to the flexors when the sleeve is worn on the right arm.

**Fig. 5.**
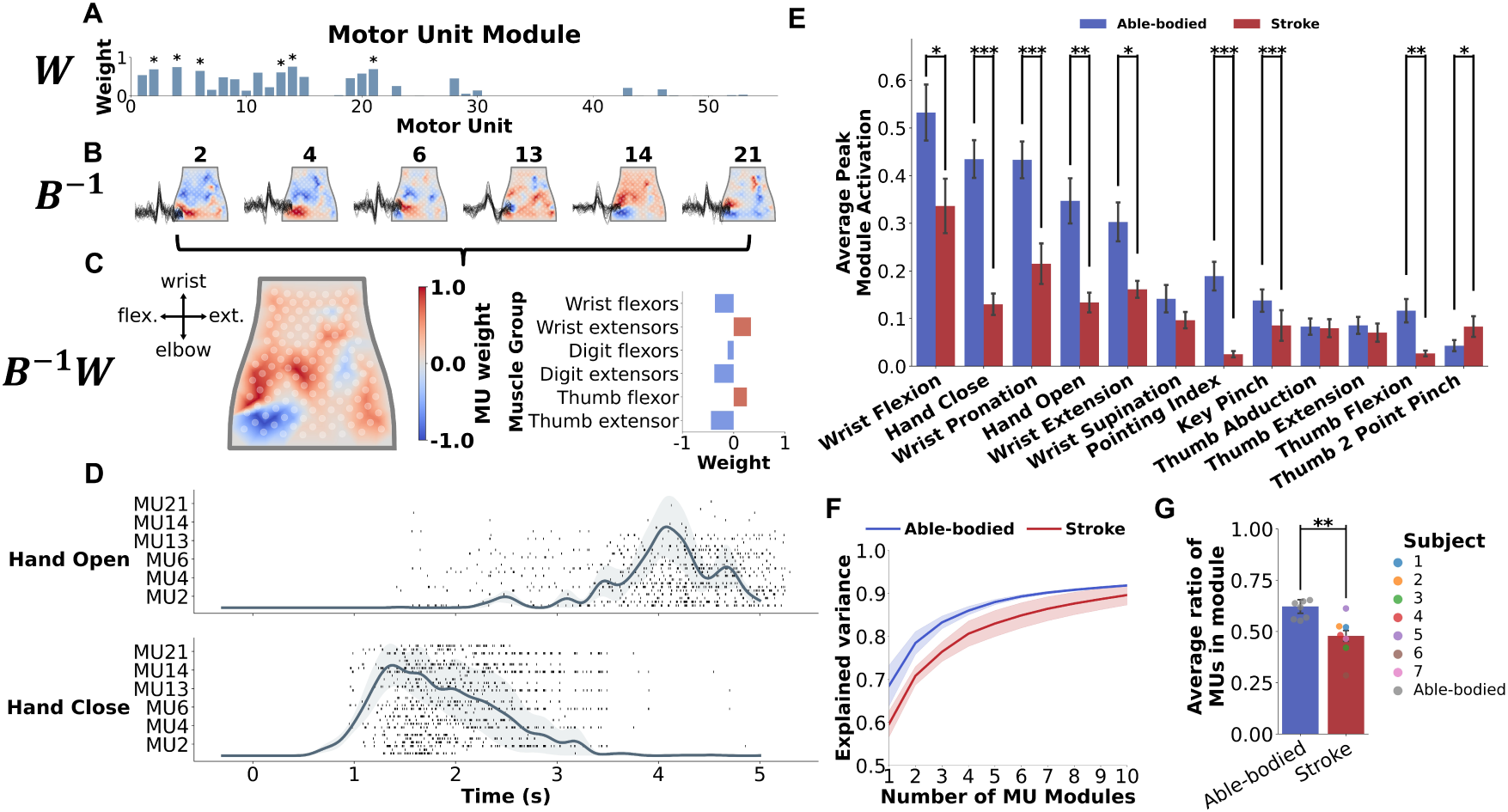
Motor unit (MU) modules within and across muscles. **A.** A single motor unit module weighting (***W*** ) to motor units decomposed using non-negative matrix factorization of motor unit firing rates. **B.** Visualization of motor units with strong weighting in module mapped to the flattened sleeve array via the inverse separation filters (***B^−^*^1^**) averaged across extensions. Spike triggered average motor unit action potential waveforms are shown on top of the heatmaps. **C.** Motor unit modules averaged across extensions and visually mapped (***B^−^*^1^*W*** ) to the sleeve array (left) and muscle groups (right). **D.** Spike train of dominant motor units in a module over five repetitions of the attempted movement. Each row shows motor unit activity during a single movement repetition. The corresponding module activation is shown on top of the spike train aligning with the active dominant motor units. **E.** Average peak module activation by movements. Able-bodied subjects had significantly higher peak module activation compared to stroke subjects for 8 of the 12 movements. **F.** Average variance accounted for (VAF) between the original and reconstructed motor unit firing rates ± standard error of the mean. Able-bodied subjects required fewer modules to reconstruct the motor unit firing rates adequately (*>* 0.85). **G.** Average ratio of the number of motor units in a module over number of total motor units decomposed. On average, able-bodied subjects had a higher percentage of motor units per module.

Figure 5D shows the average module activation *±* SEM (solid line and error shading) over time (*H*) for the five repetitions of the Hand Open and Hand Close movement attempts. The module is activated early during Hand Close, corresponding to digit flexion, whereas it is activated later in Hand Open attempts, corresponding to cocontraction of the joint while holding the hand open. The predominant motor units in the module are shown beneath the module activation. Each row within a motor unit (MU) corresponds to a single repetition of the movement attempt. The individual motor unit spiking activity aligns closely with the module activation.

To assess changes in collective motor unit activation in individuals with stroke, average peak module activation during the cue was compared between able-bodied and stroke groups (Figure 5E). Average peak activation across the most active module during a movement attempt was reduced in 8 of the 12 movements in individuals with stroke (Wrist Flexion: *p* = 0.011, Hand Close: *p* = 8.78 *×* 10*^−^*^8^, Wrist Pronation: *p* = 1.75 *×* 10*^−^*^4^, Hand Open: *p* = 1.55 *×* 10*^−^*^3^, Wrist Extension: *p* = 0.015, Pointing Index: *p* = 3.01 *×* 10*^−^*^6^, Key Pinch: *p* = 7.97 *×* 10*^−^*^4^, and Thumb Flexion: *p* = 0.003). For Thumb 2 Point Pinch only, module activation was higher in the stroke group than the able-bodied group (*p* = 0.033).

The number of modules required to reconstruct the original motor unit firing rates with at least 85% VAF varied between subjects and groups (Figure 5F). Within the stroke group, there was more variability between subjects in VAF reconstruction across module components with an average standard deviation of 0.065 *±* 0.003, with the able-bodied group at 0.029 *±* 0.011 (unpaired t-test: *p* = 0.007), highlighting the variability in hand function between participants with stroke. Additionally, more modules were required to reconstruct motor unit firing patterns in the stroke group (*n* = 7) compared to the able-bodied group (*n* = 4), suggesting potential compensatory strategies in individuals with stroke. To test this hypothesis, we evaluated the ratio of the number of motor units in a module over the total number of motor units decomposed. We found that able-bodied subjects, on average, have a greater ratio of motor units per module compared to the stroke group (unpaired t-test: *p* = 0.008), indicating that motor unit modules in able-bodied individuals tend to recruit a larger set of motor units, while stroke participants exhibit more fragmented module organization.

### 3.4 The common synaptic input to motor units reveals changes in neural drive in individuals with stroke

To assess changes in coordinated firing across frequency bands as a proxy for common synaptic input, we conducted four motor unit coherence analyses (Figure 6). We performed a global coherence analysis to assess common synaptic input across all decomposed motor units regardless of which muscle they innervated. Intra-flexor and intra-extensor coherence analyses revealed firing coordination within muscle groups. Lastly, we evaluated inter-flexor-extensor motor unit firing coherence to understand synchronized input to both muscle groups. Figure 6A-B show examples from three participants as they attempted to open their hands. In this example, there is a noticeably different firing pattern between participants, with the able-bodied subject recruiting more motor units at the beginning of the cue compared to the stroke subjects. (Figure 6A). Additionally, the spike trains reveal a reduction in firing rate for the more severe stroke participant (subject 5). Coherence analyses showed that there was an increase in synchronization of flexor motor units (non-agonist muscle) for subject 5 compared to the able-bodied and mild subjects (Figure 6B - top), whereas there was a reduction in intra-extensor motor unit coherence for the stroke participants (Figure 6B - bottom).

**Fig. 6.**
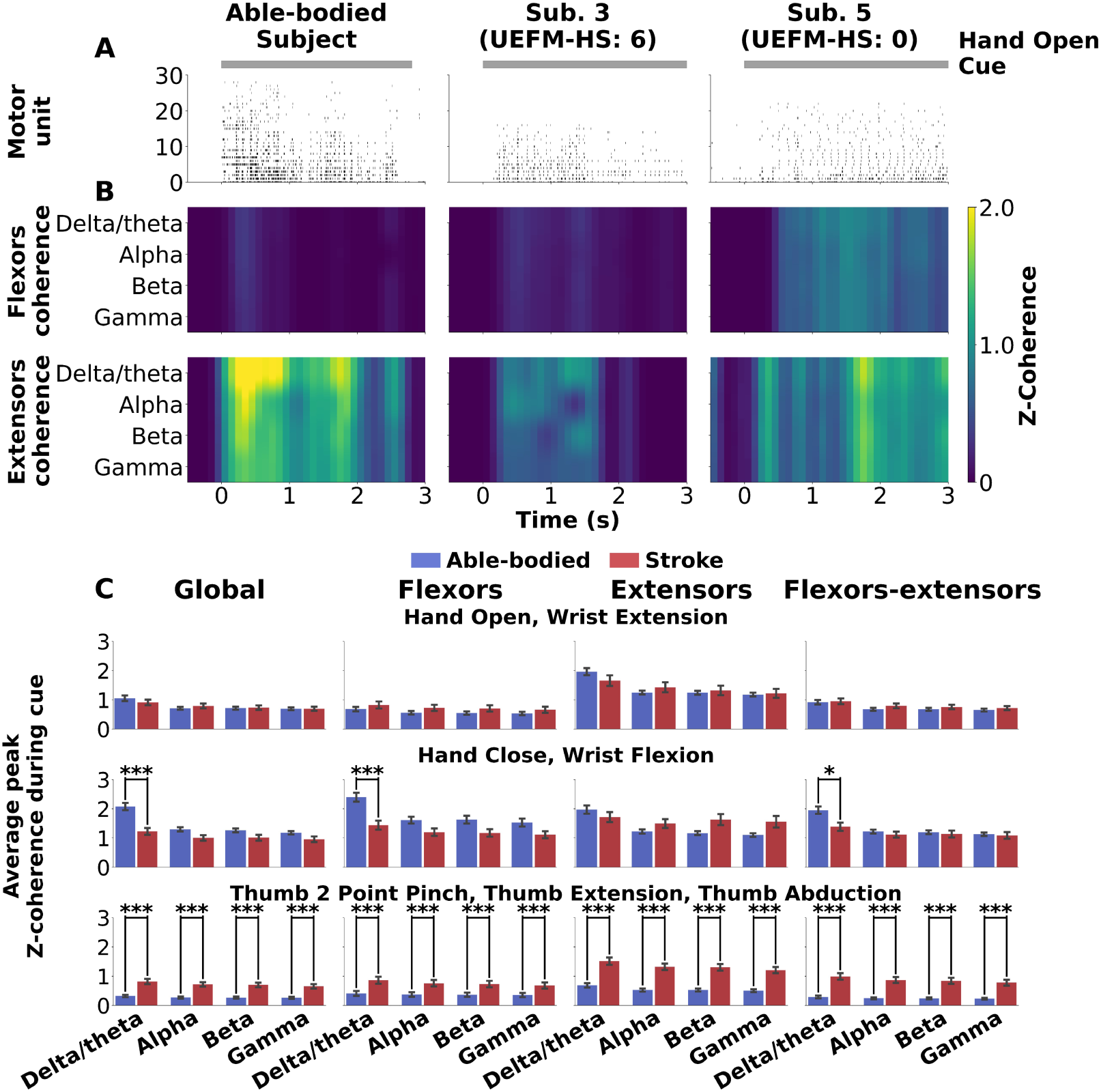
Common synaptic input to motor units within and across muscle groups. **A.** Spike train decomposed for one Hand Open repetition for an able-bodied subject, mild subject, and more severely impaired subject based on the Upper-Extremity Fugl-Meyer Hand-Subscore (UEFM-HS). **B.** Motor unit Z-coherence between randomly generated cumulative spike trains across four frequency bands (delta/theta: 0-8 *Hz*, alpha: 8-12 *Hz*, beta: 12-30 *Hz*, and gamma: 30-120 *Hz*); **top:** intra-flexors, **bottom:** intra-extensors. **C.** Average peak Z-coherence during cues across the four coherence analyses, namely global, intra-flexors, intra-extensors, and inter-flexors-extensors.

Figure 6C shows results from the four coherence analyses across a subset of movements. For extension-based movements (Hand Open and Wrist Extension), despite subject 5 having an increase in flexor motor unit coherence for Hand Open, there was no significant difference between able-bodied and stroke groups across the four analyses. However, for flexor-based movements (Hand Close and Wrist Flexion), there is a significant reduction in the global delta/theta motor unit coherence band for individuals post-stroke across all motor units (p=1.89 *×* 10*^−^*^5^), within flexor motor units (p=2.85 *×* 10*^−^*^4^), and between flexor and extensor motor units (p=0.013), while there was no significant difference within extensors. This signifies a reduction in common input related to force generation for flexion while not altering activation of the antagonist motor units.

This reduction in coherence, however, is not consistent across all movement attempts. When evaluating more dexterous movements involving the thumb, such as Thumb 2 Point Pinch, Thumb Extension, and Thumb Abduction, motor unit coherence was greater for individuals post-stroke across all frequency bands and all analyses (*p <* 0.001). This suggests there may be abnormal activation of non-target muscles when attempting to perform more dexterous movements.

### 3.5 Combined neural control features can predict functional clinical metrics

Clinical scores for individuals with stroke are shown in Table 1. The range of motor function ability is relatively large (0-7 out of 14 total points on the UEFM-HS), with one subject (subject 5) having no visible overt movement ability, to a few subjects with only mild impairment. A licensed occupational therapist evaluated each movement attempt by every participant separately with an observed movement score (Figure 1). A subset of wrist movement attempts for three individuals with stroke are shown in Figure 7A. These movements were identified to be strong predictors of motor function.

**Fig. 7.**
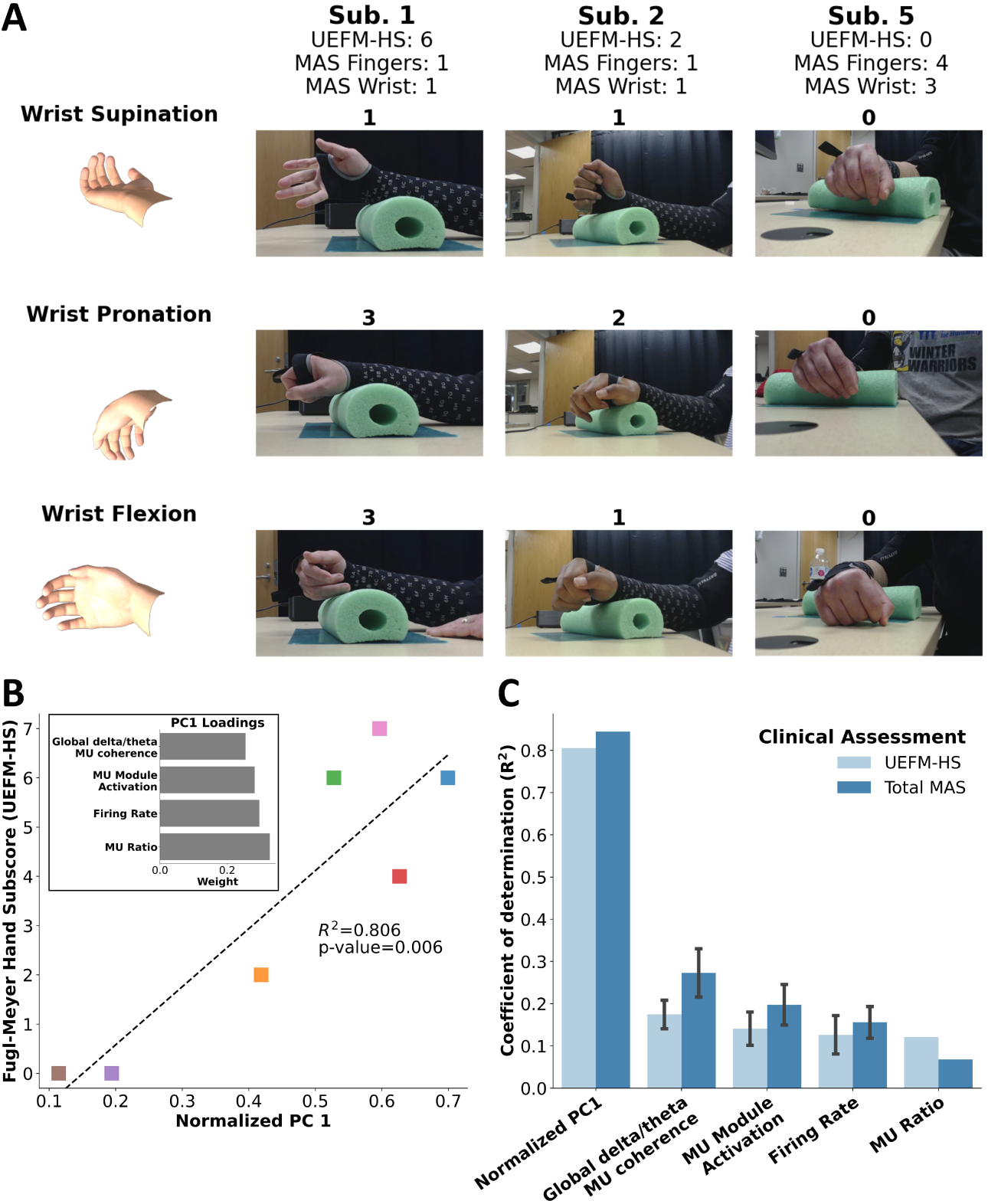
Correlating motor unit features with clinical scores. **A.** Examples of three wrist movements cued to participants and their attempts. The corresponding ARAT movement score is shown above each movement attempt per participant. **B.** Correlation of the first principal component of the combined motor unit feature dataset consisting of the three select movements and Upper- Extremity Fugl-Meyer Hand-Subscore (UEFM-HS). The combined motor unit feature dataset is made up of motor unit firing rate, motor unit module activation, motor unit ratio per module, and global delta/theta motor unit coherence. **C.** Correlation of individual features with UEFM-HS and total Modified Ashworth Scale (MAS) (MAS Fingers + MAS Wrist) using the coefficient of determination (*R*^2^) for a normalized metric.

To assess whether EMG features have the potential to identify changes in muscle activation and predict functional clinical metrics, we combined the analyzed motor unit firing characteristics in this work to create a neural control signature. Motor unit firing rate, motor unit module activation, motor unit ratio per module, and global motor unit coherence in the delta/theta band were concatenated into a single feature dataset per subject. Only global motor unit coherence was considered due to two participants with stroke not having enough muscle-specific motor units (subject 4: flexors, subject 6: extensors). Delta/theta band was prioritized due to its significantly different activation during flexion tasks when comparing individuals post-stroke and able-bodied individuals.

Principal component analysis was used to reduce this combined feature set into a singular neural control signature. The first principal component was correlated with UEFM-HS with *R*^2^ = 0.81 (*p* = 0.006) as shown in Figure 7B. Positive weights for all of the first principal component loadings indicate that as the magnitude of features increase for the select movements, hand motor function improves based on UEFM-HS. Compared to individual motor unit features, the combined signature explained motor function better than any of the other motor unit features (Figure 7C).

## 4 Discussion

This study demonstrates the potential of using a wearable HD-EMG sleeve to capture detailed motor unit activity in individuals with chronic stroke. Our findings provide insights into the neuromuscular alterations post-stroke and highlight the utility of integrating multiple motor unit properties to develop a comprehensive neural control signature that is correlated with motor function. These findings offer rehabilitation practitioners a new avenue to assess motor recovery beyond visual observation or ordinal scales. Objective neural control features, such as firing rate and motor unit coherence may help differentiate between true recovery and compensatory strategies, a key distinction when tailoring intervention plans for skill reacquisition versus substitution.

Similar to previous studies [5, 26], we found a significant reduction in motor unit firing rates in individuals with stroke during specific movements, such as wrist flexion and hand open. Additionally, we observed notable differences in motor unit recruitment and de-recruitment periods between stroke survivors and able-bodied individuals analogous to the findings by Hu et al. [21]. Stroke survivors exhibited a delayed time to reach maximum firing rates, indicating slower rate of initial motor recruitment. Furthermore, their ability to sustain muscle contractions was compromised, as evidenced by a shorter de-recruitment period. These findings suggest that stroke survivors experience both impaired initiation and maintenance of muscle contractions, reflecting broader disruptions in motor unit recruitment [5]. From a clinical perspective, reduced firing rates likely reflect impaired neuromuscular activation that contributes to functional limitations commonly observed post-stroke. These may include reduced strength, poor endurance, and difficulty with hand-based tasks such as grasping, pinching, or manipulating small objects, skills that are essential for activities of daily living and heavily targeted in occupational therapy.

In addition to changes in motor unit firing rate, we identified alterations in motor unit coupling and activation following stroke. The reduced module activation observed in 8 of the 12 movements attempted suggests a disruption in the pooled activation of motor units, which is crucial for coordinated motor control [21, 31, 65]. Fewer modules were required to reconstruct motor unit firing patterns in the able-bodied group compared to the stroke group. Additionally, the ratio of motor units per module for able-bodied individuals was greater than that of individuals post-stroke. This may indicate that stroke survivors employ compensatory motor strategies leading to the recruitment of additional modules to achieve similar tasks due to module fractionation [39], although future studies should investigate this further. These findings indicate significant changes in coordinated motor unit module recruitment which could serve as potential biomarkers of disrupted movement coordination after stroke and reveal insights into underlying motor mechanisms.

To further understand the common synaptic input to motor units, we conducted multiple coherence analyses between and within muscle groups to understand changes in motor unit inputs. We observed a reduction in coherence for gross hand and wrist movements and an increase in coherence for more dexterous hand movements involving the thumb in individuals post-stroke. This suggests abnormal activation of non-target muscles during fine movements, likely due to reliance on secondary and more grossly activating pathways (e.g., reticulospinal pathway) as a result of the damage to the corticospinal pathway that is typically responsible for more dexterous control [46]. The reticulospinal tract plays a major role in upper extremity flexion, which contributes to the upper extremity flexion synergy exhibited by individuals post-stroke who become increasingly reliant on this pathway with greater impairment to the corticospinal tract [66]. The reduction in delta/theta band coherence during flexion may be explained by the reduced neural output achieved during these tasks due to the strength of the signal. The increased coherence across all bands during more dexterous tasks may be explained by the inability to selectively activate muscles for fine motor control with a grossly activating pathway.

Our findings are further contextualized by recent work in able-bodied individuals, which showed that the dominant hand, typically more dexterous, exhibits lower coherence and thus reduced common synaptic input compared to the less dexterous non-dominant hand [45]. This supports the interpretation that increased coherence in our stroke cohort, particularly during dexterous tasks, reflects a loss of selective, independent muscle control and a shift toward more synchronized, less flexible neural drive. These results are also consistent with previous studies reporting increased coherence in the affected hand of stroke participants during highly dexterous tasks, such as isometric abduction force tracking of the index finger [4].

Examining individual motor unit features can provide insights into specific aspects of neural control of movement, but we found that a combination of features can offer a more holistic measure of motor function. When combining motor unit firing rate, motor unit module activation and ratio, and global coherence into a singular neural control signature, the first principal component of the concatenated feature set was strongly correlated with UEFM-HS (*R*^2^ = 0.81). Comparatively, Dai et al. [4] showed a weak correlation between an increase in motor unit coherence band (i.e., percentage increase of paretic compared to non-paretic) and UEFM (*R*^2^ = 0.26 *−* 0.50). Hu et al. [21] found that there was no correlation between interquartile range of motor unit size, median threshold, and interquartile range of recruitment threshold with UEFM. In a follow-up study, Hu et al. [5] achieved *R*^2^ = 0.15 when correlating the goodness of fit of the inverse power function of motor unit firing rate and UEFM. In our study, we achieved similar correlations with UEFM for individual motor unit features (Figure 7C). However, we found that combining features using an unsupervised principal component analysis leverages both information about motor unit firing activity and common inputs into an interpretable metric of neuromuscular motor function. By incorporating a user-friendly wearable system, this work demonstrates the feasibility of translating non-invasive wearable neurotechnology into clinical practice to enhance motor rehabilitation for individuals post-stroke.

While the study presents promising results, certain limitations should be acknowledged. The small sample size and use of a single data collection session may limit the generalizability of the findings. Future studies should aim to include larger, more diverse cohorts and longitudinal assessments to evaluate how these motor unit features change over time and throughout recovery. Additionally, although motor units were decomposed across multiple movements, the use of fixed separation filters may not fully capture the dynamic variability in motor unit activity that occurs with realworld functional use. Incorporating baseline isometric contractions or repeated testing sessions could provide a more robust comparison to dynamic movement attempts.

Despite these limitations, the current findings highlight the potential of a wearable HD-EMG sleeve to provide rehabilitation practitioners, including physical therapists, occupational therapists, and other neurorehabilitation specialists, with objective, quantitative insight into neural activation patterns post-stroke. Monitoring changes in motor unit firing behavior, coordination, and common drive may help clinicians better characterize impairment, track progress, and tailor interventions to address specific patterns of motor dysfunction. Integrating this technology alongside traditional clinical assessments could support more individualized and targeted rehabilitation strategies, ultimately enhancing functional outcomes and recovery trajectories for individuals with stroke.

## 5 Conclusion

This study demonstrates the utility of a wearable HD-EMG sleeve in evaluating motor unit activity in chronic stroke survivors. We observed significant reductions in motor unit firing rates and altered recruitment patterns in individuals with stroke as they attempted fine hand movements. Altered coupling and activation patterns were evident, with stroke survivors requiring more modules to reconstruct motor unit firing patterns, suggesting potential compensatory motor strategies. Coherence analyses demonstrated that stroke survivors exhibited reduced coordinated firing during gross flexion movements while showing an increased coherence during dexterous thumb movements, suggesting potential co-contraction of non-target muscles that limits the dexterous control of individuals post-stroke. To create a more encompassing metric of motor function, we combined features using an unsupervised approach into a neural control signature, which showed a strong correlation with clinical motor function scores. This combined neural control signature explained significantly more variance compared to any single motor unit metric, highlighting the strength of this approach for both research and clinical practice. These findings demonstrate the potential of a wearable HD-EMG sleeve to serve as a valuable tool to individualize treatment planning, monitor neural recovery, and guide precision-based interventions that support meaningful functional improvements in individuals post-stroke.

## Data Availability

All data produced in the present study are available upon reasonable request to the authors.

## 6 Declarations

### 6.1 Ethics approval and consent to participate

Data were collected as part of an ongoing clinical study (IRB0773, IRB 0779) being conducted at Battelle Memorial Institute that was approved by the Battelle Memorial Institute Institutional Review Board. All participants provided written informed consent before participation, in accordance with the Declaration of Helsinki.

### 6.2 Consent for publication

Consent was obtained from participants for publication of these data.

### 6.3 Availability of data and materials

Raw data were generated at Battelle Memorial Institute. Derived data and code supporting the findings of this study are available upon reasonable request. Please contact the corresponding author (Nicholas Tacca) to request access.

### 6.6 Authors’ contributions

NT developed algorithms, performed data analysis, prepared figures, and wrote the manuscript. JL provided domain knowledge expertise and supported manuscript writing. MH supported manuscript writing. BS, AB, CD, and PP provided technical support and reviewed the manuscript. The conception of the study was a collaborative effort by EM, MD, SC, and DF. EM, MD, LW, and SC carried out the data collection. LW, JP, DF, and EM provided domain expertise and co-supervised the study. All authors reviewed and approved the final version of the manuscript.

## 6.7 Acknowledgements

The authors would like to thank the broader Battelle development, quality, and management teams for their engineering work and general project support.

## 6.8 Disclaimer

The NeuroLife^®^ RECLAIM^TM^ EMG sleeve was used in the study referenced. This device has not been approved or cleared as safe or effective by FDA. This device is limited by U.S. federal law to investigational use.

