## Supplementary Information for "Leveraging neural drive to assess hand motor function in individuals with chronic stroke"

**Inclusion and Exclusion Criteria**

For physically impaired individuals, inclusion criteria address the minimum length of time since the stroke that led to the impairment. Inclusion criteria may also pertain to meeting dimensional requirements related to interacting with the system hardware (e.g., subject's arm dimensions must be such that they can appropriately don an existing electrode sleeve design). For populations with potential for cognitive impairment (e.g., stroke survivors), inclusion criteria indicating ability to follow 3-step commands and communicate verbally (e.g., at least able to provide yes/no responses with accuracy) apply. Specific Inclusion Criteria include:

1. Males and females $\geq$ 18 years old
2. Chronic stroke survivors who are at least 180 days post-stroke
3. Ability to provide appropriate consent to partake in the study
4. Ability to follow 3-step commands and deemed by an occupational therapist to have the capacity to complete required upper extremity movements
5. Ability to secure transportation to attend scheduled study sessions
6. Stroke-related hand impairment that interferes with ability to complete activities of daily living and is classified as Stage 1-6 on the hand subscale of the Chedoke McMaster Stroke Assessment

Persons with life-supporting or sustaining equipment or critical non-removeable implanted electronic devices are excluded for safety reasons since it is not known if the experimental systems would interfere with this equipment. Specific exclusion criteria include:

1. Presence of any other clinically significant medical comorbidity for which, in the judgment of the Investigator, participation in the study would pose a safety risk to the subject
2. Currently participating in physical rehabilitation (e.g., occupational or physical therapy) for stroke-related upper limb impairment
3. Co-occurring neurological condition (e.g., Parkinson's disease, Multiple Sclerosis) or other neuromuscular disorder (e.g., Carpal Tunnel Syndrome, neuropathy) that, in the judgment of the Investigator, may influence study results
4. Individuals who are immunosuppressed, have conditions that typically result in becoming immunocompromised, taking chronic steroids, or currently receiving immunosuppressive therapy
5. Individuals having or requiring any of the following: implanted pacemaker, life supporting/sustaining equipment, or critical non-removable implantable electronic devices such as an insulin pump or neurostimulator. An implanted Medtronic LINQ monitor does not meet this criterion (i.e., patients with a LINQ monitor may participate in this study).
6. Persistent pain $\geq$ 7/10 in impaired upper extremity, as measured by Numeric Pain Rating Scale (0-10)
7. Individuals whose forearm is determined to be too small or too large to fit the electrode sleeve being investigated.
8. Individuals who are pregnant or plan to get pregnant during the course of the study (self- report).

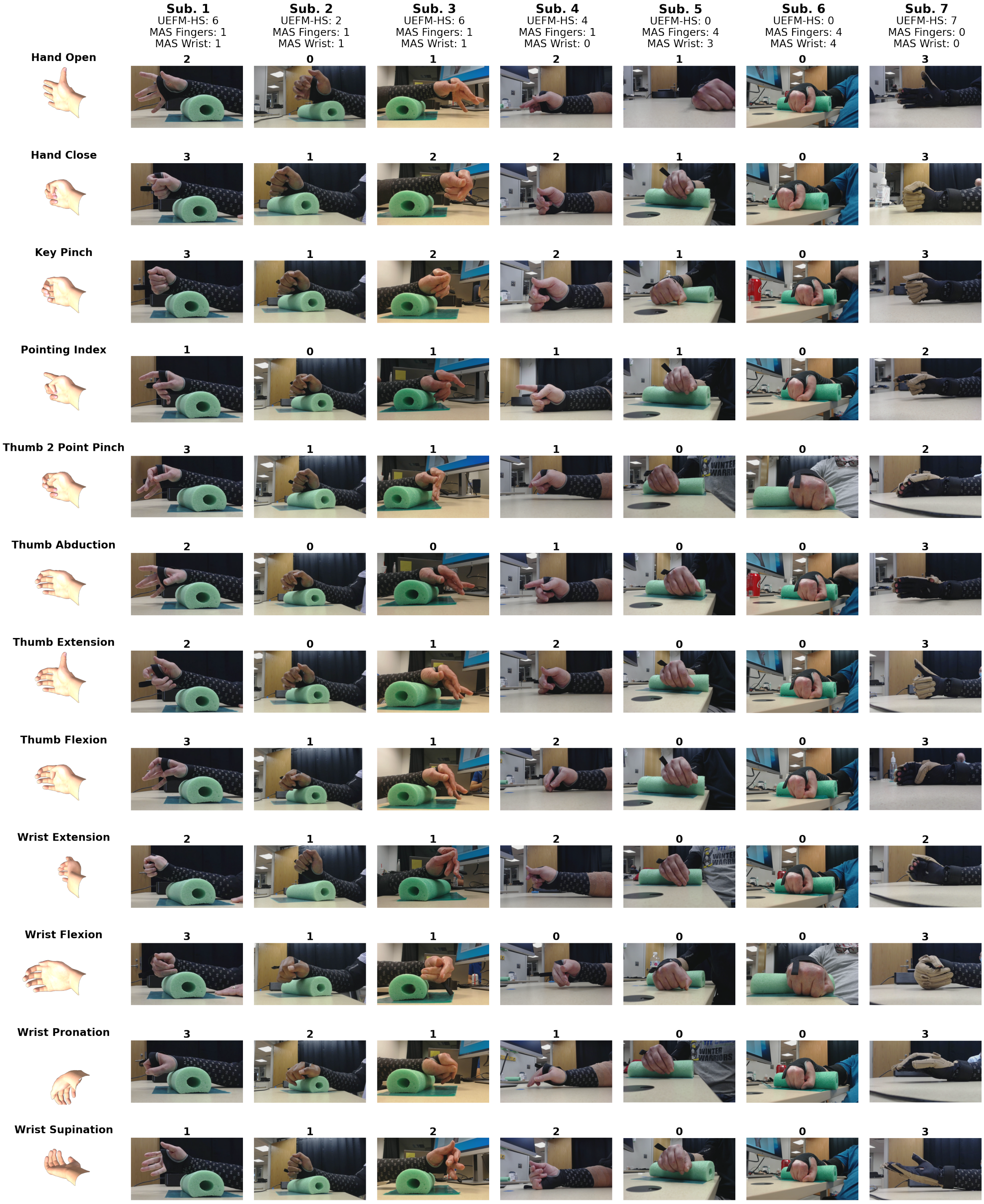


**Supplementary Figure 1 Observed movement score for individuals with stroke.** The full 12-movement dataset cues and screenshots of participant attempts. The observed movement score is above each subject's movement attempt.
